# Choroid Plexus Volume in Multiple Sclerosis can be estimated on structural MRI avoiding contrast injection

**DOI:** 10.1101/2023.05.24.23289773

**Authors:** Valentina Visani, Francesca B. Pizzini, Valerio Natale, Agnese Tamanti, Alessandra Bertoldo, Massimiliano Calabrese, Marco Castellaro

## Abstract

**Background:** Choroid Plexus (ChP) manual segmentation performance evaluation on non-contrast-enhanced MRI sequences compared to the gold-standard contrast-enhanced T1-w has never been done on a relevant cohort of subjects.

**Purpose:** To investigate whether contrast-enhancing can be avoided when performing ChP manual segmentation. To select which non-contrast-enhanced sequence between T1-w and FLAIR could be used as a contrast-enhanced T1-w surrogate. To provide a quantification of the ChP volume error that non-contrast-enhanced sequences introduce.

**Materials and methods:** Sixty-one prospective Multiple Sclerosis patients were included in the study. ChP was separately segmented on T1-w, FLAIR, and contrast-enhanced T1-w sequences. Quantitative contrast metrics between the ChP and surrounding ventricles were calculated. Quantitative segmentation metrics were obtained using gold-standard segmentation as reference. To assess the spatial agreement between non-contrast-enhanced and contrast-enhanced sequences, the segmentations were non-linearly coregistered to the standard MNI152 space and the error distribution per slice was evaluated spanning axially and coronally.

**Results:** Concerning contrast metrics, ANOVA test revealed a statistically significant main effect between the sequences (pvalue<0.01). The post-hoc t-tests revealed higher Contrast-to-Noise-Ratio and Signal-to-Noise-Ratio for contrast-enhanced sequences than others. T1-w exhibits the lower Contrast-to-Noise-Ratio while Signal-to-Noise-Ratio was comparable between FLAIR and T1-w (mean Signal-to-Noise-Ratio/Contrast-to-Noise-Ratio: contrast-enhanced T1-w=23.77/18.49, T1-w=13.73/7.44, FLAIR=13.09/10.77). The segmentation metrics revealed that non-contrast-enhanced sequences had comparable Dice. FLAIR overestimated the ChP volume while T1-w introduced a lower bias (Percentage Volume Difference FLAIR:28.02±19.02%; T1-w:3.52±12.61%). The spatial variability analysis confirmed that ChP volume depiction presents spatial differences between segmentations. FLAIR generally underperformed T1-w.

**Conclusion:** The quantitative analyses suggest that T1-w might be a good candidate as a surrogate of contrast-enhanced sequence for the ChP manual segmentation task to estimate ChP volume. On the contrary, FLAIR introduces a systematic overestimation bias.

**Summary:** To estimate the Choroid Plexus Volume with manual segmentation, contrast-enhanced T1-w can be replaced by non-contrast-enhanced T1-w because the quantified error is acceptable, while FLAIR overestimates the volume.

**Key Points:**

- T1-w MRI sequence has the lowest contrast-to-noise ratio among the available sequences but provided lower error in evaluating the Choroid Plexus Volume compared to FLAIR, both looking at spatial and overall indices (Percentage Volume Difference=3.52±12.61%). FLAIR has a higher contrast-to-noise ratio than T1-w sequence but overestimates the Choroid Plexus Volume (Percentage Volume Difference=28.02±19.02%).
- T1-w can be used as a surrogate of contrast-enhanced T1-w sequence in Choroid Plexus manual segmentation.

**Importance of the Study:** In this manuscript we present our experience concerning the use of non-contrast enhanced sequences when manually segmenting the Choroid plexus, from brain MRI. The quantification of Choroid plexus volume is becoming of great interest in recent years, however a direct comparison of gold-standard techniques based on contrast and sequences acquired without contrast agent is missing. Here we present this comparison with quantitative further analysis also on spatial pattern of manual segmentation obtained with the different sequences, suggesting a possible surrogate to the gold-standard sequence to be used in large cohort studies or to train artificial intelligence models.

## Introduction

The Choroid Plexus (ChP) is a vascular tissue located in the brain ventricular system in the four ventricles, and it forms a major part of the Blood Cerebrospinal Fluid Barrier. The main role of the ChP is the production of the majority of the Cerebrospinal Fluid (1). Moreover, ChP is a mediator of the brain clearance pathways that allow maintaining brain homeostasis (2,3), consequently, can be considered part of the glymphatic system (4). In addition, the ChP is involved in inflammatory processes, and it has been suggested to further investigate ChP role in promoting intrathecal inflammation mechanisms (5). Therefore, the functional and anatomical modification of the ChP can lead to alterations that help characterize neurodegenerative pathologies like Multiple Sclerosis (5–9), Alzheimer’s disease (2,10,11) or psychiatric disorders (12–14). The functions of the ChP have been investigated using different quantitative imaging modalities, like Diffusion Weighted Imaging (15), perfusion imaging and PET (12,16). ChP alteration can be also quantified by calculating its volume (ChPV) which has been found severely enlarged in neurological disorders (7,8,12,14). A recent study (17) has hypothesized a correlation between the inflammatory state, the ChPV, and the staging of Multiple Sclerosis, proposing the ChPV as a possible biomarker to better understand the evolution of the disease (18).

The high contrast and resolution of structural MRI have made it the natural choice to perform this segmentation task. In fact, the ChP imaging gold standard MRI technique is the T1-weighted MRI sequence enhanced with contrast injection (cT1-w) (10,19). However, cT1-w is invasive (20) and it is not routinely acquired, thus, several studies employed commonly available T1-w MRI sequences without contrast agent injection to quantitatively estimate the ChPV (21). Despite the previous uses of non-contrast enhancing T1-w sequences for the manual segmentation (MSeg) of the ChP, a quantitative assessment of the performance of MSeg of the ChP on non-contrast-enhanced sequences with respect to the gold standard cT1-w on a large cohort of patients has, to the best of our knowledge, never been done.

The aim of this work was to compare the ChP segmentations manually depicted on both cT1-w, T1-w and FLAIR (Fig. 1) to determine whether non-contrast-enhanced sequences are accurate enough to be used in quantitative studies. Moreover, we will provide an estimation of the error in ChPV quantification using non-contrast-enhanced sequences when compared to cT1-w sequences, suggesting whether the contrast injection might be avoided for ChPV quantification.

**Figure 1:**
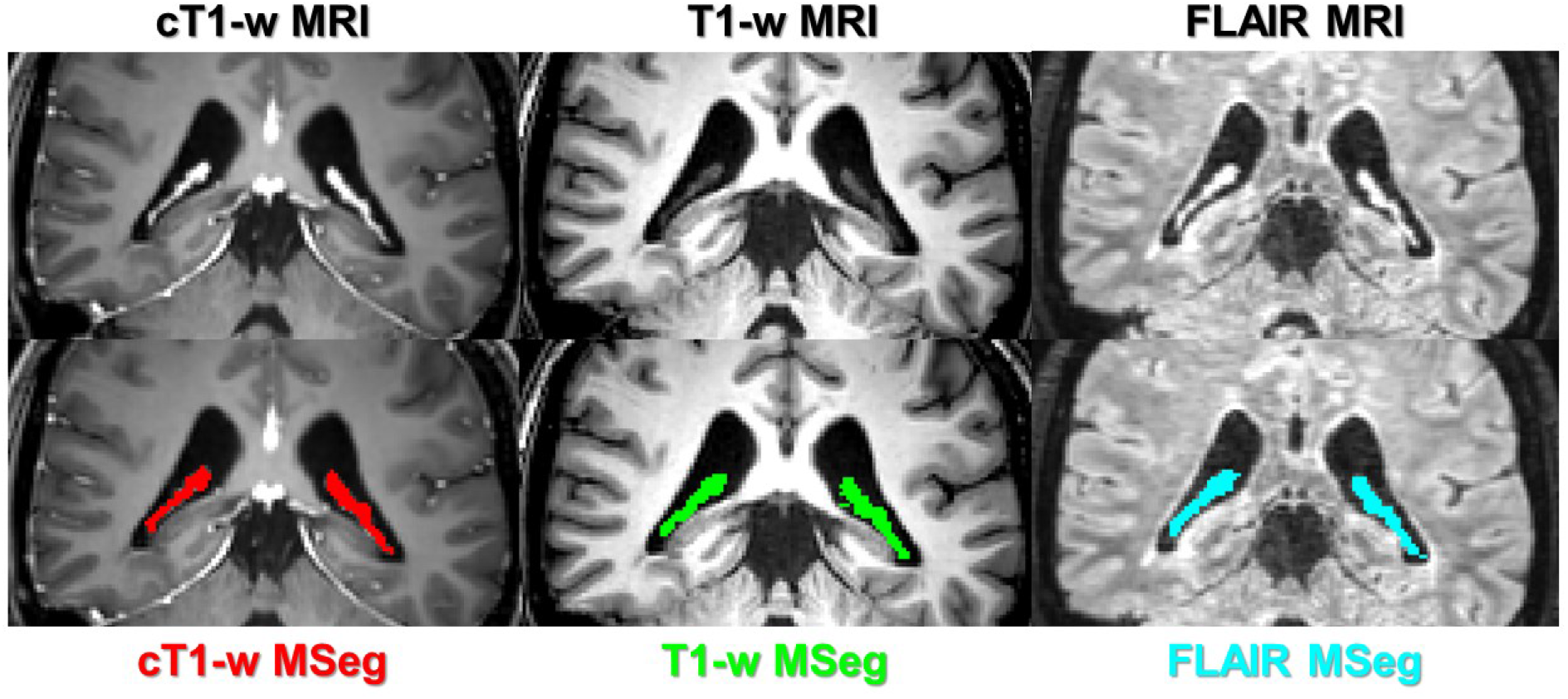
Coronal view of Choroid Plexus of a representative patient - first row: from left to right, cT1-w, T1-w and FLAIR MRI; second row: from left to right, previous images with overlapped, respectively, cT1-w manual segmentation (MSeg) in red, T1-w MSeg in green and FLAIR MSeg in blue.

## Materials and Methods

The study workflow is reported in Figure 2 and was composed by the following steps.

**Figure 2:**
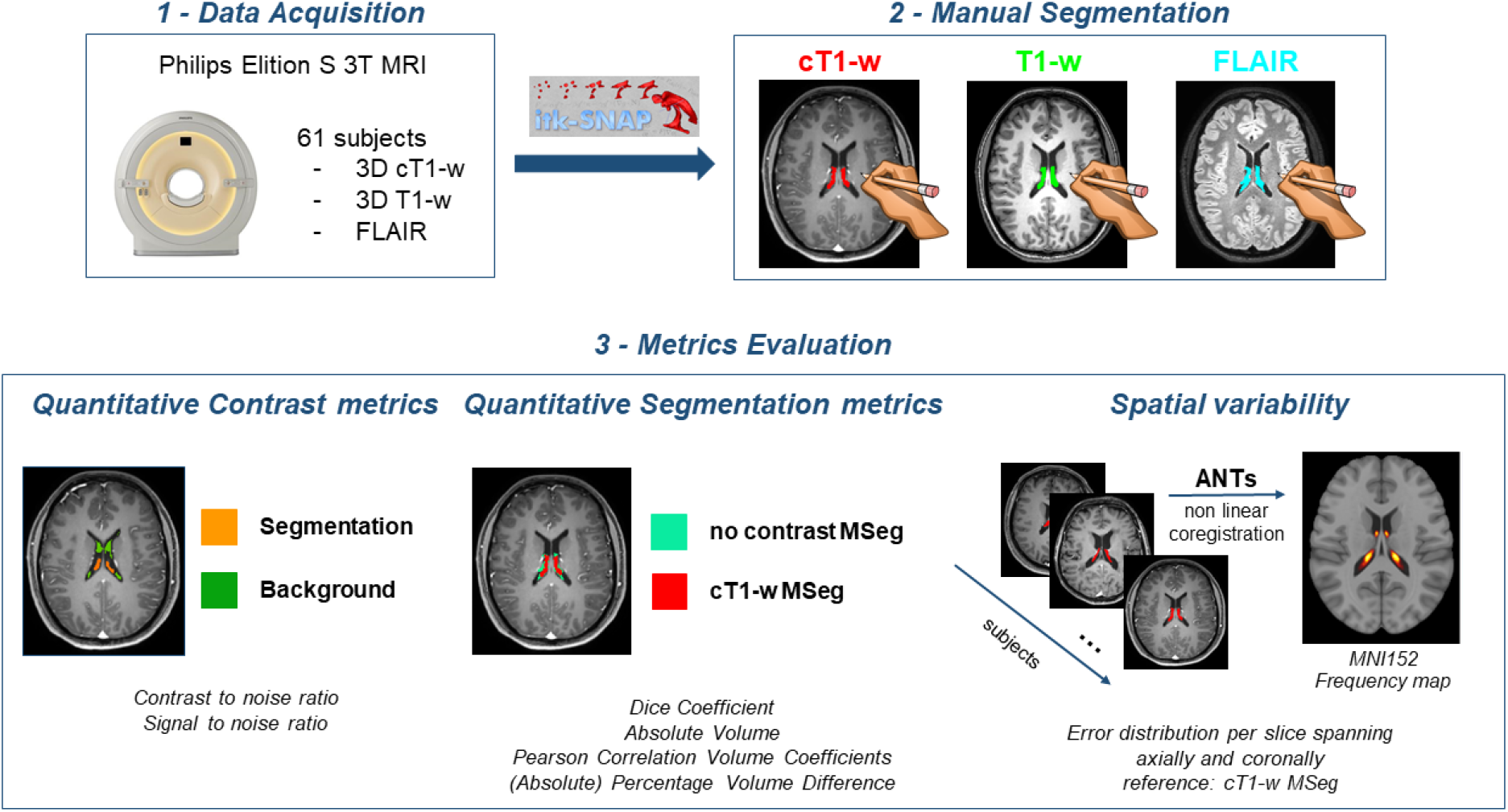
Workflow of the study. 1 - Data acquisition: see Methods-Data acquisition for details; 2 - Manual Segmentation: performed by neuroradiologists on ITK-snap for each available sequence for each subject (see Methods-Manual segmentation for details); 3 - Metrics Evaluation: analyses performed in the study (see Methods-Quantitative contrast metrics - Quantitative segmentation metrics - Spatial variability for details). ANTs: Advanced Normalization Toolbox.

### Data acquisition

All patients gave their written informed consent prior to participating in the study. All procedures were performed in accordance with the Declaration of Helsinki (2008) and the study protocol was approved by the local Ethical Committee. Sixty-one Relapsing-Remitting Multiple Sclerosis patients (age 39.9±9.5 years) acquired prospectively between November 2020 and October 2021 were included. Images were acquired on a Philips Elition-S 3T MRI scanner, equipped with a 32 channels head coil with this protocol: 3D T1-w (MPRAGE: CS-SENSE factor: 4; TE/TR: 3.8/8.5ms; FA: 8°; 1×1×1 mm); FLAIR (3D Turbo Spin Echo – CS-SENSE factor: 5; TE/TR: 376/8000 ms; TI: 2356 ms); cT1-w with same parameters as 3D T1-w after 10 minutes from injection of contrast (Gadovist 1.0 mmol/mL).

### Manual segmentation

The manual segmentation procedure was done in a common space on cT1-w and FLAIR after coregistering both sequences to the T1-w. A junior neuroradiologist depicted the MSegs of the ChP in the two lateral ventricles (21) on each sequence and for each patient using ITK-snap (22) and a senior neuroradiologist confirmed the segmentations. Patients were randomized and each sequence was segmented separately. The order of segmentation was T1-w, FLAIR, and cT1-w to limit the rater bias towards the cT1-w sequence.

### Quantitative contrast metrics

We investigated the contrast-to-noise ratio (CNR) and the signal-to-noise ratio (SNR) between the ChP and the background region (lateral ventricles) (Tab. 1). The ChP region-of-interest was obtained from the MSegs. The background region-of-interest was obtained as follows. Firstly, a raw ventricles segmentation was extracted from the FreeSurfer (v7.1.1) pipeline (23). The raw ventricle mask was refined by excluding the union of the ChP masks obtained for each sequence (T1-w, FLAIR, and cT1-w). Lastly, an erosion operation (spherical kernel of 2 mm) was applied to include only background voxels.

**Table 1:** Metrics formulations. ROI-Choroid Plexus (ChP) region; background-ventricles region; ref-reference segmentation (cT1-w); segm - compared segmentation; TP - true positive, voxel correctly classified as ChP (1) in the segm compared to ref; FP – false positive, voxel wrongly classified as ChP in the segm compared to the ref; FN – false negative, voxel wrongly classified as background in the segm compared to the ref.

| Quantitative Contrast metrics |  |
| --- | --- |
| *Contrast to Noise Ratio* | $\frac{\text{mean}(ROI_{\text{values}}) - \text{mean}(\text{background}_{\text{values}})}{\text{std}(\text{background}_{\text{values}})}$ |
| *Signal to Noise Ratio* | $\frac{\text{mean}(ROI_{\text{values}})}{\text{std}(\text{background}_{\text{values}})}$ |
| Quantitative Segmentation metrics |  |
| *Dice Coefficient* | $\text{Dice}(\text{ref}, \text{segm}) = \frac{2 * TP}{2 * TP + FP + FN}$ |
| *Percentage Volume Difference* | $100 * \frac{(\text{Volume}_{\text{segm}} - \text{Volume}_{\text{ref}})}{\text{Volume}_{\text{ref}}} [\%]$ |
| Spatial variability metrics |  |
| *Error distribution per slice* | $100 * \frac{(\text{VoxelSlice}(i)_{\text{segm}} - \text{VoxelSlice}(i)_{\text{ref}})}{\text{Volume}_{\text{ref}}} [\%]$ |

### Quantitative segmentation metrics

To quantitatively investigate the differences between the MSegs obtained from the available sequences, we calculated for each subject, using the cT1-w MSeg as reference: the Dice Coefficient; the Absolute Volume; the Pearson Correlation Volume Coefficients; the Percentage Volume Difference (ΔVol%); the Absolute Percentage Volume Difference (|ΔVol%|) (Tab. 1).

### Spatial variability

To assess the spatial variability of each sequence, we non-linearly co-registered, using the Advanced Normalization Toolbox (24), each subject to the MNI Talairach ICBM 152 2009c Nonlinear Symmetric template (MNI152) (25). We evaluated the error distribution per slice (Tab. 1) spanning axially and coronally to provide quantitative metrics of agreement between the non-contrast-enhanced sequences and the cT1w sequence. Lastly, we constructed the frequency maps reporting for each voxel the probability of being ChP for each MSegs.

### Statistics

One-way ANOVA and post-hoc t-test (significance level α=0.05) between the available sequences were performed for both SNR and CNR values. Regarding quantitative segmentation metrics, one-way ANOVA and post-hoc t-test (α=0.05) were performed for Absolute Volume metric between all available MSeg. One-sample t-test (α=0.05) was performed for ΔVol% metric for both no-contrast-enhanced MSegs. Two-sample t-test (α=0.05) was performed for both ΔVol% and |ΔVol%| metrics between two no-contrast-enhanced MSegs.

## Results

Table 2 reports the results for both the quantitative contrast and segmentation metrics.

**Table 2:** Results of Quantitative Contrast metrics and Quantitative Segmentation metrics presented in form mean ± standard deviation. Quantitative Contrast metrics: contrast-to-noise ratio and signal-to-noise ratio between ChP MSeg obtained from each sequence and a reference region obtained excluding the ChP from the ventricles (see Methods-Quantitative contrast metrics for details); Quantitative Segmentation metrics: Dice Coefficient, Absolute Volume, (Absolute) Percentage Volume Difference and Pearson’ Volume Correlation Analysis between MSegs obtained from non-contrast enhanced sequences (T1-w and FLAIR MSeg) and the reference cT1-w MSeg. ChP: Choroid Plexus; MSeg: Manual Segmentation; cT1-w: T1-weighted MRI sequence enhanced with contrast injection.

| Metrics | T1-w | cT1-w | FLAIR |
| --- | --- | --- | --- |
| ***Quantitative Contrast metrics*** |  |  |  |
| *Signal to Noise Ratio* | 13.73±4.89 | 23.77±12.02 | 13.09±6.15 |
| *Contrast to Noise Ratio* | 7.44±2.87 | 18.49±9.57 | 10.77±5.38 |
| ***Quantitative Segmentation metrics*** |  |  |  |
| *Dice Coefficient* | 0.67±0.05 | - | 0.68±0.05 |
| *Absolute Volume [mm<sup>3</sup>]* | 3073±563 | 2984±506 | 3787±679 |
| *Percentage Volume Difference [%]* | 3.52±12.61 | - | 28.02±19.02 |
| *Absolute Percentage Volume Difference [%]* | 10.57±7.60 | - | 28.43±18.40 |
| *Pearson' Correlation* | 0.77 | - | 0.67 |

For both the contrast metrics (SNR and CNR), the ANOVA revealed a statistically significant main effect between the tested sequences (SNR: p_value_<0.01; CNR: p_value_<0.01). As shown in Figure 3, the post-hoc t-tests between each sequence combination showed a statistically significant higher SNR and CNR for cT1-w when compared to both T1-w and FLAIR (SNR/CNR: cT1-w=23.77/18.49, T1-w=13.73/7.44, FLAIR=13.09/10.77). FLAIR had statistically significant higher CNR when compared to T1-w (p_value_<0.01). T-test between T1-w and FLAIR on SNR did not show significant differences (p_value_=0.10).

**Figure 3:**
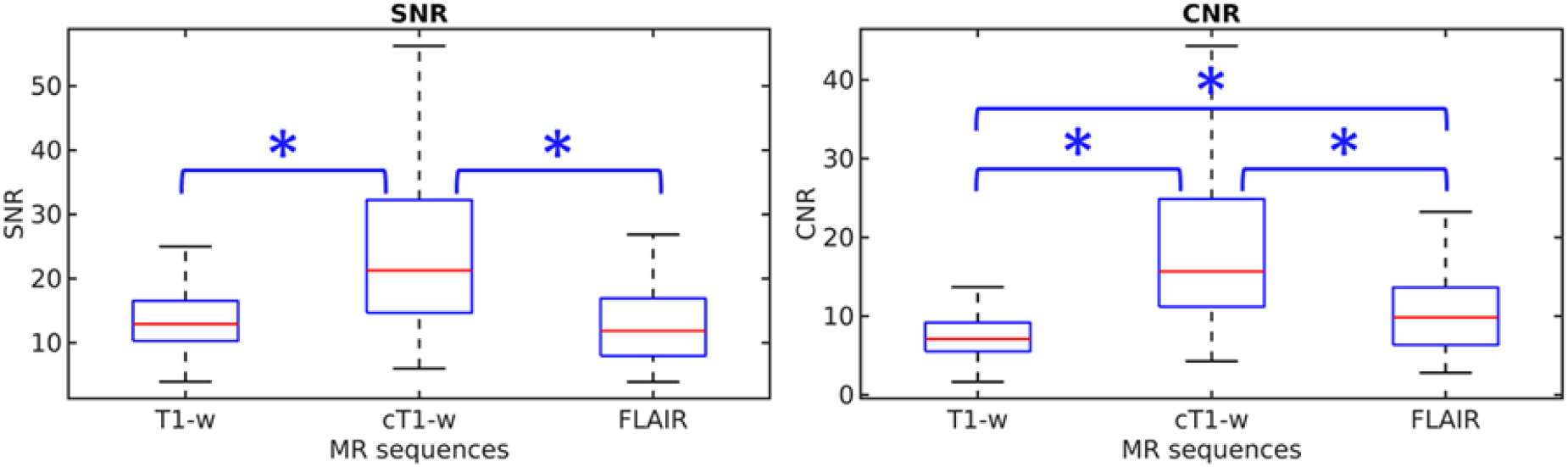
Quantitative contrast metrics - Boxplot of signal-to-noise ratio (SNR) and contrast-to-noise ratio (CNR) between the Choroid Plexus and the ventricles for the three compared MRI sequences T1-w, FLAIR, and contrast-enhanced T1-w. The asterisk indicates that the metrics between the two groups are statistically different (pvalue<0.05).

The pair-wise correlation analysis between each MSeg obtained from each sequence reported a significant positive correlation between the volume of cT1-w MSeg and both non-contrast-enhanced sequences (T1-w vs cT1-w: 0.77; FLAIR vs cT1-w: 0.67). One-way ANOVA revealed a statistically significant main effect between tested MSegs (p_value_<0.01). FLAIR had statistically higher Absolute Volume when compared to both T1-w and cT1-w MSegs (p_value_<0.01), while there were no statistically significant differences between T1-w and cT1-w MSegs volumes (p_value_=0.36). T1-w MSeg provides lower |ΔVol%| (10.57±7.60%) compared to FLAIR MSeg (28.43±18.40%), that overestimates ChPV for every subject (ΔVol% T1-w: 3.52±12.61%; FLAIR: 28.02±19.02%) (Fig. 4). Statistical tests on |ΔVol%| and ΔVol% confirmed the two no-contrast-enhanced MSegs have statistical differences (p_value_<0.01). One-sample t-test revealed on ΔVol% T1-w is not normally distributed with zero-mean with a p_value_=0.03. The MSegs obtained from non-enhanced sequences have similar Dice Coefficients (T1-w vs cT1-w: 0.67; FLAIR vs cT1-w: 0.68) (Tab. 2).

**Figure 4:**
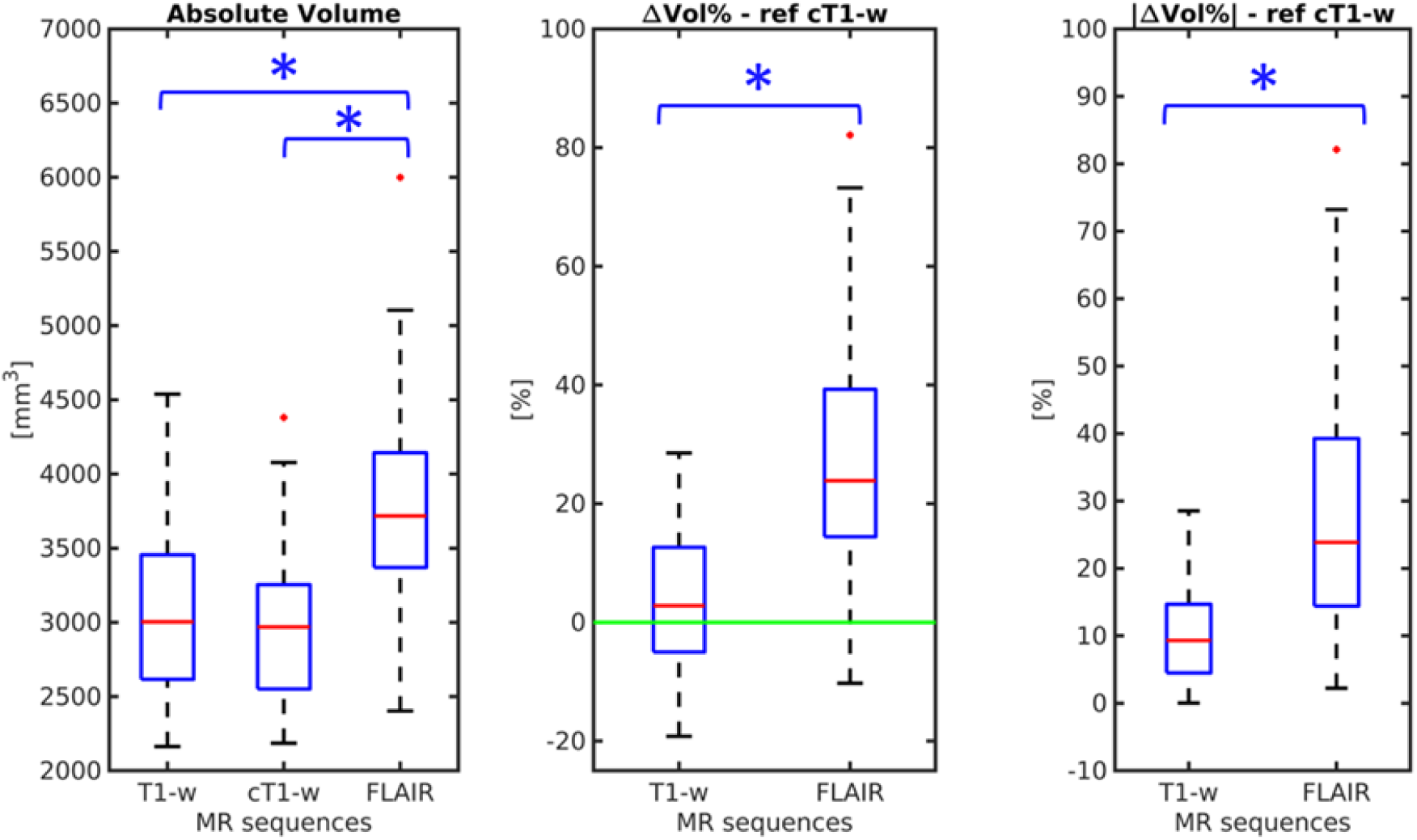
Quantitative segmentation metrics - from left to right: boxplot of the Absolute Volume of the Manual segmentations (MSegs); boxplot of the Percentage Volume Difference (ΔVol%) for T1-w and FLAIR MSegs (reference cT1-w); boxplot of the Absolute Percentage Volume Difference (|ΔVol%|) for T1-w and FLAIR MSegs (reference cT1-w). The asterisk indicates statistically significant differences between groups (pvalue<0.05).

The spatial variability analysis we conducted in the MNI space provided an error distribution per slice that confirms that ChPV presents spatial differences between MSegs (Fig. 5). Axial-wisely, FLAIR was more in agreement than T1-w with the reference cT1-w only in the temporal horn near the anterior boundary of the hippocampus and the lateral ventricles (MNI152 z-coordinate = -12). The T1-w sequence was better aligned to the cT1-w in the other portions of the ChP while the FLAIR sequence tends to suggest a larger volume in the trigonum ventriculi (MNI 152 z-coordinate = 10). In the body of the lateral ventricles, near the fornix (MNI152 z-coordinate = 18) it was detected a less accurate agreement between non-contrast and contrast-enhanced sequences (mean error: T1-w>0.7%, FLAIR>1.2%), probably due to the tiny dimension of the ChP that run parallel to the fornix up to the proximal portion of the anterior horn of the ventricles. Analyzing the ChP coronal-wise, the T1-w was in better agreement with the cT1-w sequence in the majority of the ChP. In the atrium of the lateral ventricles (MNI152 y-coordinate = -42) Figure 5 shows a great variability of both non-contrast-enhanced sequences, however, the T1-w bias was lower than FLAIR. The most posterior portion of the ChP, located in the occipital horns of the lateral ventricles (MNI y-coordinate = -35), is often overestimated by the FLAIR sequence (error=0.8%). Lastly, the ChP ends near the anterior part of the fornix (MNI y-coordinate = -7), where we measured a comparable error between FLAIR and T1-w sequences (error=0.5%).

**Figure 5:**
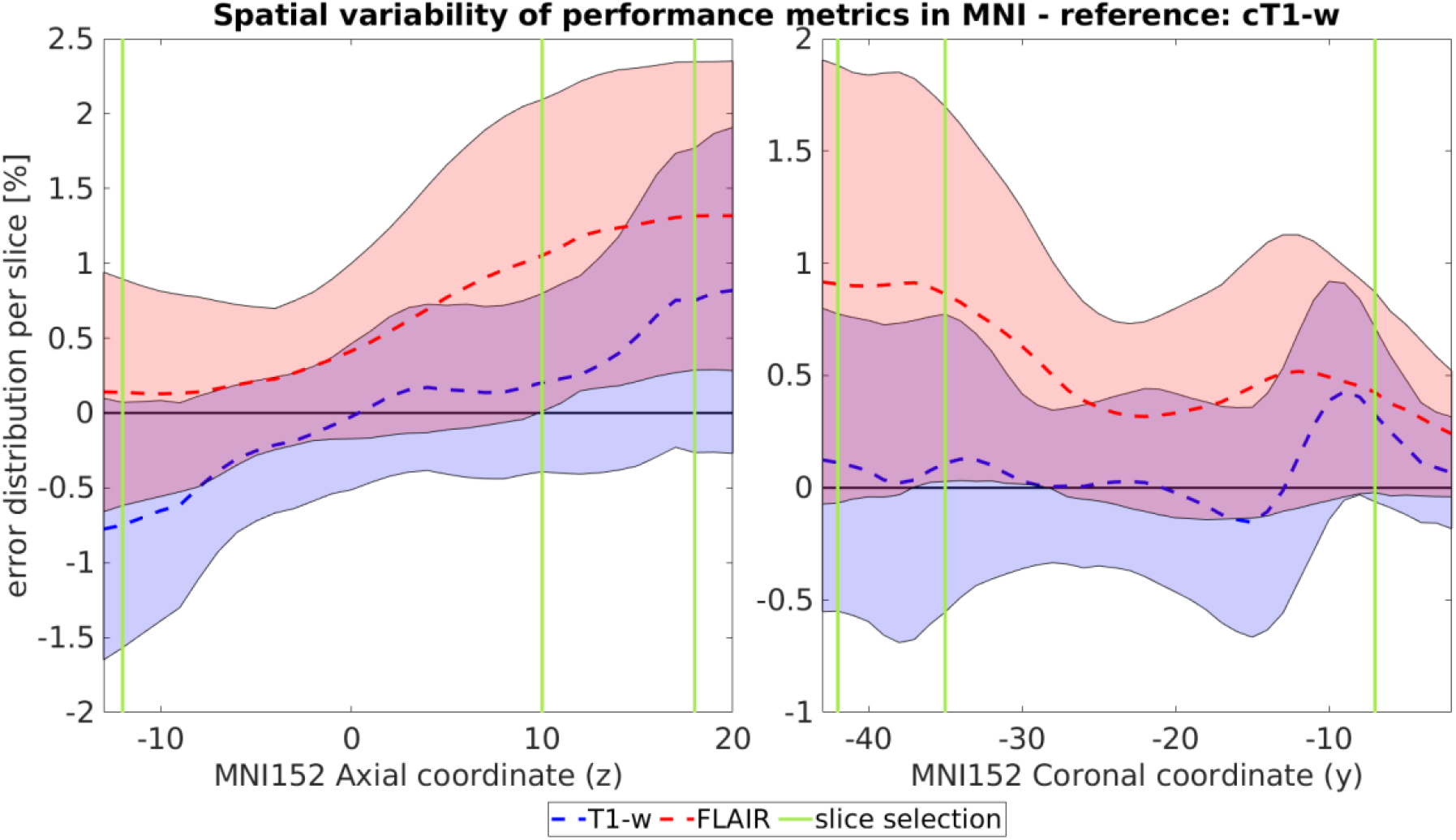
Slice-wise evaluation of performance metrics in MNI Talairach ICBM 152 2009c Nonlinear Symmetric template (MNI152) coordinate system. The graph represents the error distribution per slice between no contrast manual segmentations (FLAIR in red, T1-w in blue) and the reference cT1-w computed axial-wise along the z-coordinate (on the left) and coronal-wise along the y-coordinate (on the right) of the MNI152 coordinate system. The vertical green lines highlight three representative axial (z=-12: temporal horn near the anterior boundary of the hippocampus, z=10: trigonum ventriculi, z=18: body of the lateral ventricles near the fornix) and coronal (y=-7: atrium of the lateral ventricles, y=-42: occipital horns of the lateral ventricles, y=-35: anterior portion of the fornix) slices reported in Figure 6.

Figure 6 shows the frequency maps of the MSegs overlapped to the MNI152 atlas. The frequency map of the cT1-w MSeg is reported as reference, while for the T1-w and FLAIR MSegs it is reported the difference between the frequency map and the reference. Both axially and coronally, FLAIR sequence tends to overestimate the ChPV, while the T1-w behaves more similar to cT1-w.

**Figure 6:**
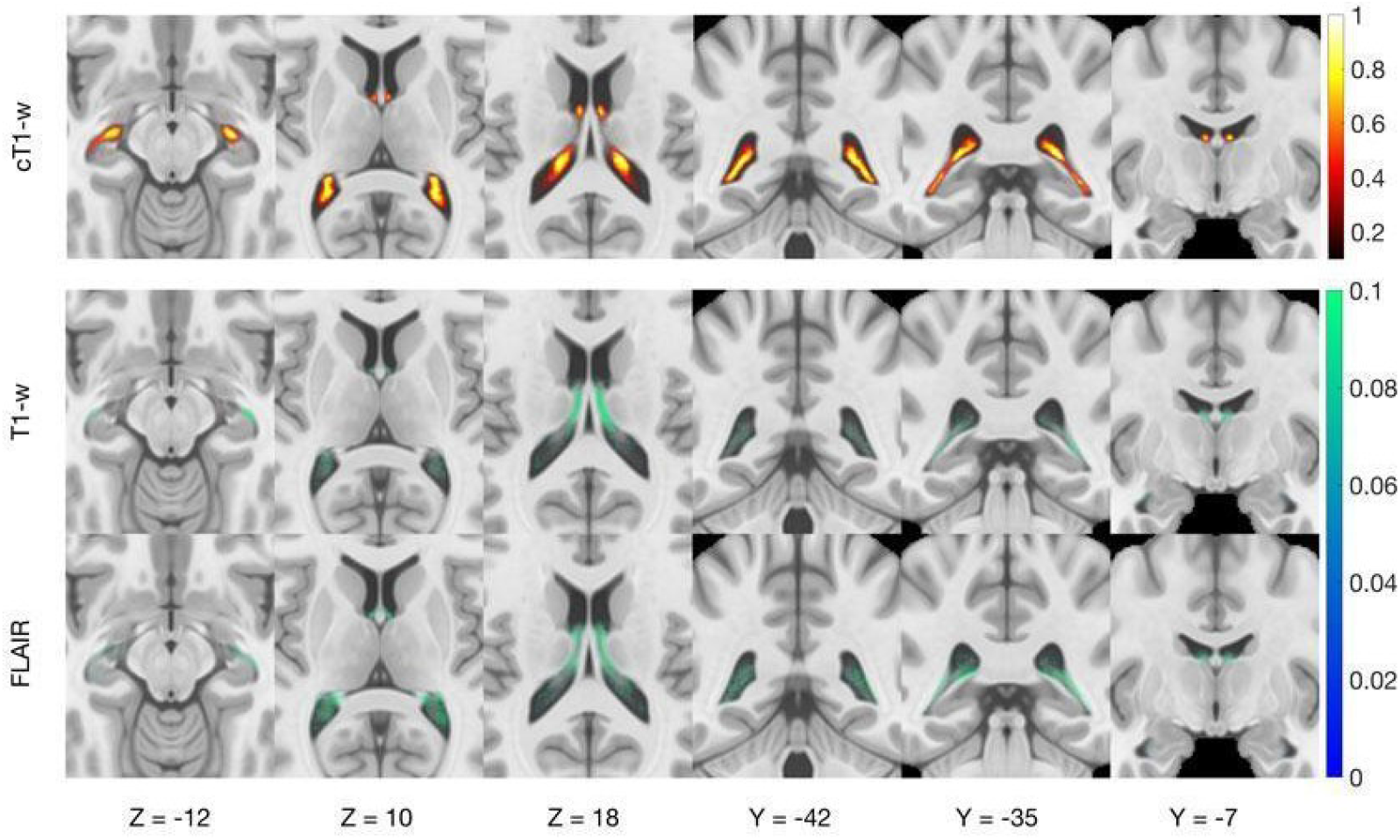
Probability frequency maps of manual segmentations (MSegs) overlapped to the MNI Talairach ICBM 152 2009c Nonlinear Symmetric template (MNI152) - From left to right, three axial views (z=-12: temporal horn near the anterior boundary of the hippocampus, z=10: trigonum ventriculi, z=18: body of the lateral ventricles near the fornix) and three coronal views (y=-7: atrium of the lateral ventricles, y=-42: occipital horns of the lateral ventricles, y=-35: anterior portion of the fornix); coordinates reported are centered to the origin of the MNI space. The first row represents the frequency map of the cT1-w MSeg for representative slices overlapped to the MNI152 template. The second and third row, respectively show the difference between the frequency map of MSegs depicted on non-contrast enhanced sequences (T1-w and FLAIR MSeg) and the MSeg obtained from the cT1-w sequence overlapped to the MNI152.

## Discussion

To our knowledge this is the first study that quantitatively evaluates the manual segmentation of the ChP obtained from different sequences, both in terms of intrinsic contrast, overlapping metrics and spatial variability in a population of Multiple Sclerosis patients. The aim of our work was to investigate whether the use of cT1-w sequences, considered the gold standard for ChP imaging, is necessary to quantitatively estimate the ChPV and which sequence between T1-w and FLAIR can be considered the best alternative.

When comparing contrast metrics calculated between ChP and the lateral ventricles, among the available sequences, we expected that cT1-w and FLAIR could provide better visualization features than T1-w sequences (Fig. 1). CNR and SNR confirms that cT1-w sequence is the most suitable sequence to visually inspect and depict the ChP and that FLAIR seemed its best alternative.

The segmentation metric analysis showed a good agreement between both FLAIR and T1-w sequence with the reference cT1-w. The Dice Coefficient alone in our case was not informative to detect the best candidate to substitute cT1-w. Nevertheless, more than exploring segmentation overlap, we are interested in the quantification of the ChPV. ChPV obtained from the FLAIR sequence is usually greater than one obtained with cT1-w images, as clearly shown in Figure 4, while T1-w does not provide a specific trend of bias. Moreover, T1-w commits lower systematic errors and lower error variability. This indication was also confirmed by Pearson’s correlation analysis.

Regarding spatial variability of segmentations in MNI space, FLAIR was more in agreement than T1-w with the reference cT1-w only axial-wise in the temporal horn near the head of the hippocampus and of the lateral ventricles probably due to the greater CNR of the ChP guaranteed by the FLAIR sequence that in this tiny space could potentially improve the segmentation performance. For other portions of the ChP, the intrinsic blurring of the FLAIR image when compared with the T1-w brings to an overestimate of the ChPV.

## Conclusion

The quantitative analyses we conducted suggest that T1-w might be the better candidate as a surrogate of cT1-w for the ChP MSeg task to estimate ChPV with respect to FLAIR sequences. However, when segmenting the ChP, it might be helpful to use both T1-w and FLAIR in the anterior portion of the temporal horn of the lateral ventricles. On the contrary, T1-w is to be preferred in the other ChP portions due to the fixed overestimation bias introduced by the FLAIR. Moreover, the high variability we encountered in several portion of the ChP might highlight the possibility of restricting the ChP volume segmentation, when employing non-contrast-enhanced sequences, to the central part of the ChP, for example excluding regions near the anterior temporal horn or near the fornix.

## Data Availability

All data produced in the present study are available upon reasonable request to the authors

